# Protocol for a Rapid Scoping Review of Evidence of Outdoor Transmission of COVID-19

**DOI:** 10.1101/2020.08.07.20170373

**Authors:** Mike Weed, Abby Foad

## Abstract

The COVID-19 pandemic is both a global health crisis, and a civic emergency for national governments, including the UK. As countries across the world loosen their lockdown restrictions, the assumption is generally made that the risk of COVID-19 transmission is lower outdoors, and this assumption has shaped decisions about what activities can re-commence, the circumstances in which they should re-commence, and the conditions under which they should re-commence. This is important for events and activities that generate mass gatherings, particularly mass participation sports events such as running, but also events in other sectors such as concerts, carnivals and festivals.

This paper sets out a protocol for a rapid scoping review of evidence of incidents of outdoor transmission of COVID-19, including the settings, environments and circumstances of such transmission, and their comparative prevalence to incidents of indoor transmission.

Its purpose is to inform discussions about the recommencement of activities that generate mass gatherings.

## Introduction

Following 213 global deaths and 9,800 infections, on 30^th^ January 2020 the World Health Organisation categorised COVID-19 as a Public Health Emergency of International Concern (PHEIC), and five weeks later, on 11^th^ March, as a pandemic, at which point 118,000 cases and 4,291 deaths in 114 countries had been reported (Ghebreyesus, 2020). The first cluster of COVID-19 cases were recorded in Wuhan, China on 21^st^ December 2019, and the first death on 11^th^ January 2020. In the UK, the first domestically contracted case was recorded on 28^th^ February, and the first death on 5^th^ March. A global spread across Europe and North and South America meant that countries in every continent around the world, faced with a virus for which there was no vaccine and no treatment, implemented lockdown measures to deal with a global pandemic that had resulted in over 700,000 recorded deaths worldwide by 1^st^ August 2020 (Worldometer, 2020).

Following reductions in the spread of the virus as a result of these periods of lockdown, many countries are now developing and implementing plans to cautiously open up their societies and economies. Most of Europe has loosened lockdown measures and is now putting in place mitigation measures to live with the virus, including deciding which activities can re-commence, and which cannot. In the UK, after six weeks of lockdown, the government published a COVID-19 recovery strategy on 11^th^ May, with an update for further expansion published on 24^th^ July (HM Government, 2020a).

One of the assumptions in the UK recovery plan, and those of many other countries, is that the risk of transmission of the virus is lower outdoors (HM Government, 2020a). This assumption is central in informing decisions about which parts of the economy and society, and what activities, can re-commence, including decisions about the mitigation measures required. One area in which this is of particular importance is physical activity and sports, and particularly mass participation events such as running. Following government guidelines issued on 10^th^ July (HM Government, 2020b) it has been possible for mass participation running events to take place, and some have done so under principles drawing on government guidelines issued by UK Athletics (UK Athletics, 2020). These principles require additional mitigation measures to ensure some social distancing, including but not limited to, staggered wave starts, and strategies to limit pre-event gathering (such as not having on-site briefings, registration and announcements), and to accelerate post-event dispersal. However, for many event hosts, the issue of the risk of transmission of COVID-19 outdoors remains paramount, and this paper sets out a protocol for a rapid scoping review to explore evidence of such outdoor transmission. The findings of the review will also be relevant to events in other sectors that generate mass gatherings, such as concerts, carnivals and festivals.

## Review Scope

The review will seek, evaluate and analyse evidence of incidents of outdoor transmission of COVID-19, the settings and environments of such transmission, and, where available, all relevant circumstances, including, but not limited to, temperature, wind conditions, social crowding or distancing, and the existence or otherwise of any COVID-19 mitigation measures. The review will also seek, evaluate and analyse evidence of the prevalence of outdoor transmission compared to indoor transmission, and evidence of the impact of high profile mass gatherings, both immediately before (e.g. Champions League soccer matches) and during (e.g. Black Lives Matter protests) lockdowns.

The review will not seek, evaluate or analyse evidence relating to the science of outdoor transmission. As such, it will be a review of evidence of *whether* outdoor transmission has taken place, not of *how* outdoor transmission takes place.

The review has been designed to be undertaken rapidly in 15 days. Therefore, a key additional purpose of the review will be to assess whether a further extended more detailed and comprehensive review would capture wider and more extensive evidence. It will also consider what new research could be undertaken to generate further evidence in a timely manner.

## Review Protocol

The initial search will be undertaken in Google Scholar, using the search string < “COVID 19” outdoor transmission >. Experimental searches suggest that “COVID 19” with a space within quotation marks is the most effective search term, and that adding alternatives for COVID 19 (e.g. SARS-CoV 2, Coronavirus) adds little to the efficacy of the search. Similarly, adding alternatives for “outdoor”, such as “outside” or “open air”, also does not appear to improve search efficacy.

Search results will initially be reviewed using the article title and preview text containing search terms returned by Google Scholar to evaluate whether a returned paper refers to outdoor transmission. If it appears it does, the full text of the paper will be searched for the word “outdoor” and the relevant passages of text reviewed to establish whether any evidence, or references to other sources of evidence, of outdoor transmission is included. Chains of sources referred to in the paper will be pursued to the original source or sources of evidence of outdoor transmission, which will be included for evaluation and analysis. Sources that provide opinion, summation or only onward reference to evidence of outdoor transmission will not be included. Given the rapidly emerging evidence relating to COVID-19, neither peer-review status nor the outlet in which the source is published, will be used as inclusion/exclusion criteria.

It is expected that evidence of outdoor transmission will be limited, and while it is likely to be referred to in many sources, sources containing actual evidence are likely to be limited. For this reason, the initial search, ordered by Google Scholar’s relevance function, will be limited to the first 100 sources returned. It is assumed that the search will be saturated at this point, and no new sources of evidence will be added. However, if the last 20 returns (81-100) do add further sources of evidence, then the search will be extended to the next 20 sources until the point is reached that the last 20 sources do not add any additional sources of evidence.

The search will then be repeated, ordered by Google Scholar’s date added function. This will ensure that new sources of evidence are not being overlooked. The same protocol as described above will be followed in relation to establishing the relevance of the returns, reference mining, and establishing the saturation point for the search.

In addition to the Google Scholar search, the papers and evidence sources considered by the UK government’s Scientific Advisory Group for Emergencies (SAGE), and its feeder groups, including the Scientific Pandemic Influenza Group on Modelling (SPI-M) and the New and Emerging Respiratory Virus Threats Advisory Group (NERVTAG), will be searched for evidence and discussion of outdoor transmission. Finally, specific searches of the research of known authors researching COVID-19 transmission will be undertaken.

As evidence of incidents of outdoor transmission of COVID-19 is expected to be limited, inclusion criteria will relate only to relevance – there will be no inclusion/exclusion criteria related to quality or proxies for quality. However, quality will be evaluated in the analysis and synthesis of the included sources. The product of this analysis will be a critical *narrative synthesis* (Pope & Mays, 2006) which, whilst describing and synthesising evidence in substantive terms, will also highlight potential weaknesses in returned evidence, and the caveats about conclusions that can be drawn, throughout the narrative.

MW will review and evaluate sources for inclusion on the basis of relevance, and will write the narrative synthesis, including an embedded evaluation of evidence quality. AF will review and validate the inclusion decisions, and the evaluation of evidence quality embedded in the narrative synthesis.

## Purpose

The purpose of the review is to inform discussions about the re-commencement of activities that will generate mass gatherings. Primarily, this will focus on physical activity and sport events, but it will also consider events in other sectors such as concerts, carnivals and festivals. The narrative presentation of the results of the review will reflect this purpose

## Data Availability

This is a protocol for a Rapid Scoping Review of Evidence. The data comprises the sources that will be returned in the review, all of which will be referenced and available for public scrutiny

